# Comparison of seven commercial SARS-CoV-2 rapid Point-of-Care Antigen tests

**DOI:** 10.1101/2020.11.12.20230292

**Authors:** Victor M. Corman, Verena Claudia Haage, Tobias Bleicker, Marie Luisa Schmidt, Barbara Mühlemann, Marta Zuchowski, Wendy Karen Jó Lei, Patricia Tscheak, Elisabeth Möncke-Buchner, Marcel A. Müller, Andi Krumbholz, Jan Felix Drexler, Christian Drosten

**Author notes:** Address correspondence to: Christian Drosten, Charite-Universitätsmedizin Berlin, Institute of Virology, Charitéplatz 1, 10117 Berlin Germany.

## Abstract

**Background:** Antigen point of care tests (AgPOCT) can accelerate SARS-CoV-2 testing. As first AgPOCT are becoming available, there is a growing interest in their utility and performance.

**Methods:** Here we compare AgPOCT products by seven suppliers: the Abbott Panbio™ COVID-19 Ag Rapid Test; the RapiGEN BIOCREDIT COVID-19 Ag; the Healgen® Coronavirus Ag Rapid Test Cassette (Swab); the Coris Bioconcept Covid.19 Ag Respi-Strip; the R-Biopharm RIDA®QUICK SARS-CoV-2 Antigen; the NAL von minden NADAL COVID19-Ag Test; and the Roche/SD Biosensor SARS-CoV Rapid Antigen Test. Tests were evaluated on recombinant nucleoprotein, cultured endemic and emerging coronaviruses, stored clinical samples with known SARS-CoV-2 viral loads (n=138), stored samples from patients with respiratory agents other than SARS-CoV-2 (n=100), as well as self-sampled swabs from healthy volunteers (n=35).

**Findings:** Limits of detection in six of seven tested products ranged between 2.08 × 10^6^ and 2.88 × 10^7^ copies per swab, the outlier at 1.58 × 10^10^ copies per swab. Specificities ranged between 98.53% and 100% in five products, with two outliers at 94.85% and 88.24%. False positive results were not associated with any specific respiratory agent. As some of the tested AgPOCT were early production lots, the observed issues with specificity are unlikely to persist.

**Interpretation:** The sensitivity range of most AgPOCT overlaps with viral load figures typically observed during the first week of symptoms, which marks the infectious period in the majority patients. AgPOCTs with a limit of detection that approximates the virus concentration above which patients are infectious may enable shortcuts in decision-making in various areas of healthcare and public health.

## Background

The ongoing SARS-CoV-2 pandemic challenges public health systems worldwide. In absence of effective vaccines or drugs, virus detection by RT-PCR has been widely adopted to enable nonpharmaceutical interventions based on case finding and contact tracing. Because of its superior sensitivity and specificity, RT-PCR is the gold standard for SARS-CoV-2 detection (1).

RT-PCR is a laboratory-based procedure that requires sophisticated equipment, trained personnel, as well as logistics for sample shipment and results communication. Timeliness of results is critical for the control of onward transmission due to the concentration of viral shedding around the time of symptoms (2). The widespread limitation of timely laboratory results is aggravated by the increasing demand for RT-PCR tests certified for *in-vitro* diagnostic application, creating supply bottlenecks and shortenings of overall testing capacity in many countries (3).

Antigen detection is usually inferior to RT-PCR in terms of sensitivity and specificity (4, 5). Nevertheless, the possibility to perform point of care testing can provide essential information when it is needed, even if in some situations the obtained information has to be amended by an RT-PCR result obtained at a later point. As first industry-manufactured antigen point of care test (AgPOCT) devices are becoming available, there is a growing interest in their performance with particular respect to sensitivity and overall specificity, two essential parameters that can guide decisions over fields of application (6). Because of the intense but short-lived nature of SARS-CoV-2 shedding from the upper respiratory tract, the clinical validation of AgPOCT requires great attention to the timing of infection in studied subjects (7, 8). If subjects are tested late in the course of infection, such as in the second week after onset of symptoms, incongruences between RT-PCR and AgPOCT will cause an apparently low clinical sensitivity for AgPOCT that is not necessarily relevant when using these tests to diagnose early acute infections. From a practical perspective, knowledge of the analytical-rather than clinical sensitivity of AgPOCT may be sufficient to judge their utility in various fields of application, as compared to the well-established RT-PCR as a reference method (9).

Here we aimed to compare seven available AgPOCT devices against an established RT-PCR assay (10) by conducting a single-center evaluation in a laboratory setting. Evaluation of analytical sensitivity relied on recombinant SARS-CoV-2 nucleoprotein, SARS-CoV-2 cell culture supernatants, as well as stored clinical samples with established SARS-CoV viral loads. Specificity was evaluated on cell culture supernatants containing endemic and emerging human Coronaviruses, clinical samples that earlier tested positive for respiratory pathogens, as well as fresh nasopharyngeal self-swabs of healthy subjects.

## Material and methods

### Clinical samples

All stored specimens were taken for routine diagnostic testing with no extra procedures required for the study. Specimens were stored in phosphate-buffered saline (PBS) or universal transport medium (Copan UTM™) at −20°C. Respiratory samples for specificity testing were obtained during 2019 from patients hospitalized at Charité medical center and tested by the NxTAG® Respiratory Pathogen Panel (Luminex). SARS-CoV-2 positive samples were collected between March and October 2020 and tested and quantified by the SARS-CoV-2 E-gene assay as published previously (10, 11). RNA was extracted from clinical samples by using the MagNA Pure 96 system (Roche). The viral RNA extraction was performed using 100µl of sample, eluted in 100µl. Viral RNA of human coronaviruses (CoVs) other than SARS-CoV-2 was quantified by real-time RT-PCR using specific *in vitro* transcribed RNA standards (10, 12, 13). Virus RNA concentrations are given as copies per mL.

### SARS-CoV-2 negative healthy subjects

Healthy volunteers were employees of the institute of virology, between 22 and 61 years of age (median, 34.7 years). All subjects received instructions as well as material to conduct self-testing with all AgPOCT at one point of time. All testing was done under supervision of trained personal. Of note, most manufacturers do not list self-test in their instructions for use. However, in recent study, self-sampling was shown to be a reliable alternative to professional nasopharyngeal swabs for AgPOCT (14). All manufacturers’ instructions were exactly followed during self-sampling.

### AgPOCT testing

For the evaluation of the AgPOCTs, 50µl of stored respiratory samples (swab resuspended in 1-3 mL of phosphate-buffered saline or universal transport medium) were mixed with sample buffer volume as specified in the manufacturers’ instructions. Results in the form of a band on immunochromatography paper were scored independently by two persons. In case of discrepant evaluations, a third person was consulted to reach a final decision. In case of test failure indicated by absence of a visible positive control band, the test procedure was repeated on the same sample. All SARS-CoV-2 RNA negative samples that showed a false-positive result in POCTs were retested.

### Recombinant SARS-CoV-2 nucleoprotein (SARS-CoV-2-N)

The coding sequence of the SARS-CoV-2 nucleoprotein was amplified, purified and cloned into the expression vector pET151/D-TOPO (Thermofisher Scientific). E. coli Bl21 (DE3) cells were transformed with the pET151/D-TOPO-SARS-CoV-2 N plasmid. Proteinpurification was performed by affinity chromatography under native conditions as described previously with minor modifications (15). A second purification step was included using heparine sepharose columns. N protein was eluted with a NaCl gradient. For analytical sensitivity experiments SARS-CoV-2-N protein was diluted in PBS and 50 µl of each dilution were applied to each test. Three replicates per test were performed.

### Cell culture samples

Cell culture supernatants containing all endemic human coronaviruses (HCoV)⍰229E, ⍰NL63, ⍰OC43 and ⍰HKU1 as well as MERS-CoV, SARS-CoV, and SARS-CoV-2 were tested in duplicates. Viral RNAs were extracted from cell culture supernatants by the viral RNA mini kit (Qiagen) according to the manufacturer’s instructions. RNA concentration in all samples was determined by specific real-time RT-PCR and *in vitro*-transcribed RNA standards designed for absolute quantification of viral load. In the case of SARS-CoV-2 additional quantification was done by plaque titration (11).

### Statistical analysis

Logistic regression analyses were run using the PyMC3 package in Python (16). The logistic regression model was implemented as follows:

y ∼ Bernoulli(θ)

θ = logistic(alpha + beta * X)

alpha ∼ Normal(0, 15)

beta ∼ Normal(0, 15)

Where X is the observed log_10_ SARS-CoV-2 RNA / mL, and y is the AgPOCT result. Models were run for 25000 iterations with 5000 tuning steps using the automatically assigned No-U-Turn sampler and an acceptance rate of 0.95. Models were assessed for convergence using the Gelman Rubin statistic and visualization of posterior traces. Posterior predictive distributions were used to assess model fit.

## Ethical statement

The use of stored clinical samples for validation of diagnostic methods without person-related data is covered by section 25 of the Berlin hospital law and does not require ethical or legal clearance. The ethical committee has been notified of the study and acknowledged receipt under file number EA1/369/20. The testing of employees is part of an ongoing study on SARS-CoV-2 infection in employees under Charité ethical review board file number EA1/068/20.

## Results

### Analytical Sensitivity

Initial comparisons of analytical sensitivity relied on purified bacterially-expressed viral nucleocapsid protein, the target protein of all assays. Protein concentrations between 5 and 25 ng/mL were detectable by most assays, corresponding to 250 to 1250 ng of protein per 50 µl sample volume (**Table 1**). To confirm these figures on viral protein, we tested cell culture supernatants from SARS-CoV-2-infected Vero cells at defined concentrations of infectious (plaque-forming) units (PFU) of virus. Almost all AgPOCT reliably detected ca. 44 PFU of virus per assay (**Table 1**). The assays by manufacturers I, III, V, and VII detected as little as 4.4 PFU of virus per test. The assay by manufacturer II was considerably less sensitive in detecting recombinant protein as well as virus.

**Table 1.** Outcome of testing by using serial dilutions of recombinant SARS-CoV-2 nucleoprotein and SARS-CoV-2 cell culture supernatant (triplicates). Protein and virus were diluted in PBS. 50µl was used for testing; PFU, plaque-forming unit.

|  |  | AgPOCT assay<sup>a</sup> |  |  |  |  |  |  |
| --- | --- | --- | --- | --- | --- | --- | --- | --- |
|  |  | I | II | III | IV | V | VI | VII |
| Recombinant N-protein Concentration [ng/mL] | 1,000 | 3 | 3 | 3 | 3 | 3 | 3 | 3 |
|  | 250 | 3 | 3 | 3 | 3 | 3 | 3 | 3 |
|  | 50 | 3 | 0 | 3 | 3 | 3 | 3 | 3 |
|  | 25 | 3 | 0 | 3 | 3 | 3 | 3 | 3 |
|  | 10 | 3 | 0 | 3 | 0 | 3 | 3 | 3 |
|  | 5 | 2 | 0 | 3 | 0 | 3 | 2 | 3 |
|  | 2.5 | 0 | 0 | 3 | 0 | 3 | 0 | 0 |
| SARS-CoV-2 [PFU/mL] | 8,800 | 3 | 3 | 3 | 3 | 3 | 3 | 3 |
|  | 880 | 3 | 2 | 3 | 3 | 3 | 3 | 3 |
|  | 88 | 3 | 0 | 3 | 0 | 3 | 1 | 3 |
|  | 8,8 | 0 | 0 | 0 | 0 | 0 | 0 | 0 |
|  | 0,88 | 0 | 0 | 0 | 0 | 0 | 0 | 0 |

<sup>a</sup>I: Abbott Panbio™ COVID-19 Ag Rapid Test; II. RapiGEN BIOCREDIT COVID-19 Ag; III: Healgen® Coronavirus Ag Rapid Test Cassette (Swab); IV Coris Bioconcept Covid.19 Ag Respi-Strip; V: R-Biopharm RIDA®QUICK SARS-CoV-2 Antigen; VI NAL von minden; NADAL COVID19-Ag Test; VII: Roche/SD Biosensor SARS-CoV Rapid Antigen Test

### Analytical sensitivity using clinical samples

To determine the analytical sensitivity in clinical samples, we used stored swabs obtained in universal transport medium (Copan UTM™) or without any medium. Dry swabs were suspended in phosphate-buffered saline and all swab suspensions were tested by RT-PCR as described (10). Of each suspension, 50 µl were introduced into the recommended volume of lysis reagent for each AgPOCT.

It should be noted that this procedure introduces a pre-dilution step (ca. 1:20) not normally applied in AgPOCT protocols, resulting in a loss of sensitivity as opposed to RT-PCR. On the contrary, the swabs used for this study are standard-gauge flocked swabs that are not provided with AgPOCT. The swabs provided with AgPOCT consist of the same material but are considerably thinner and thus carry less sample volume. To estimate the relative sample input in the present procedure, we inserted standard flocked swabs as well as the swabs included in AgPOCT kits in a solution of 50% sucrose and determined the relative sample volume contained in each swab by weighing. The resulting relative sample volume carried on AgPOCT swabs was ca. 40% (range, ca. 10-90%) of that in standard-gauge swabs. Taking the above-mentioned pre-dilution into account, this results in an approximately 8-fold lesser sample input in AgPOCT in the present study, as opposed to direct application as per manufacturer’s instructions. This factor should be accounted for when directly comparing against RT-PCR sensitivity in the following. It should be noted that the piece-to-piece variability of swabs in some supplier’s AgPOCT assays is considerable.

A total of 138 SARS-CoV-2 RT-PCR positive samples were tested (**Figure 1A**). Median virus load was 2.49 × 10^6^ (range: 1.88 × 10^4^ - 2.75 × 10^9^) copies per mL of swab suspension. Depending on initial testing and available volume per clinical sample, up to 115 clinical samples per assay were used to evaluate AgPOCT assays (**Figure 1B**). Only 45 samples were used for the assay by manufacturer II, which detected only 4 of 45 samples correctly, each of these four samples containing more than 2 ×10^8^ RNA copies/mL, leading us to terminate further sensitivity testing for this product. The distribution of test samples across all AgPOCT products is shown in **Figure 1B**.

**Figure 1.**
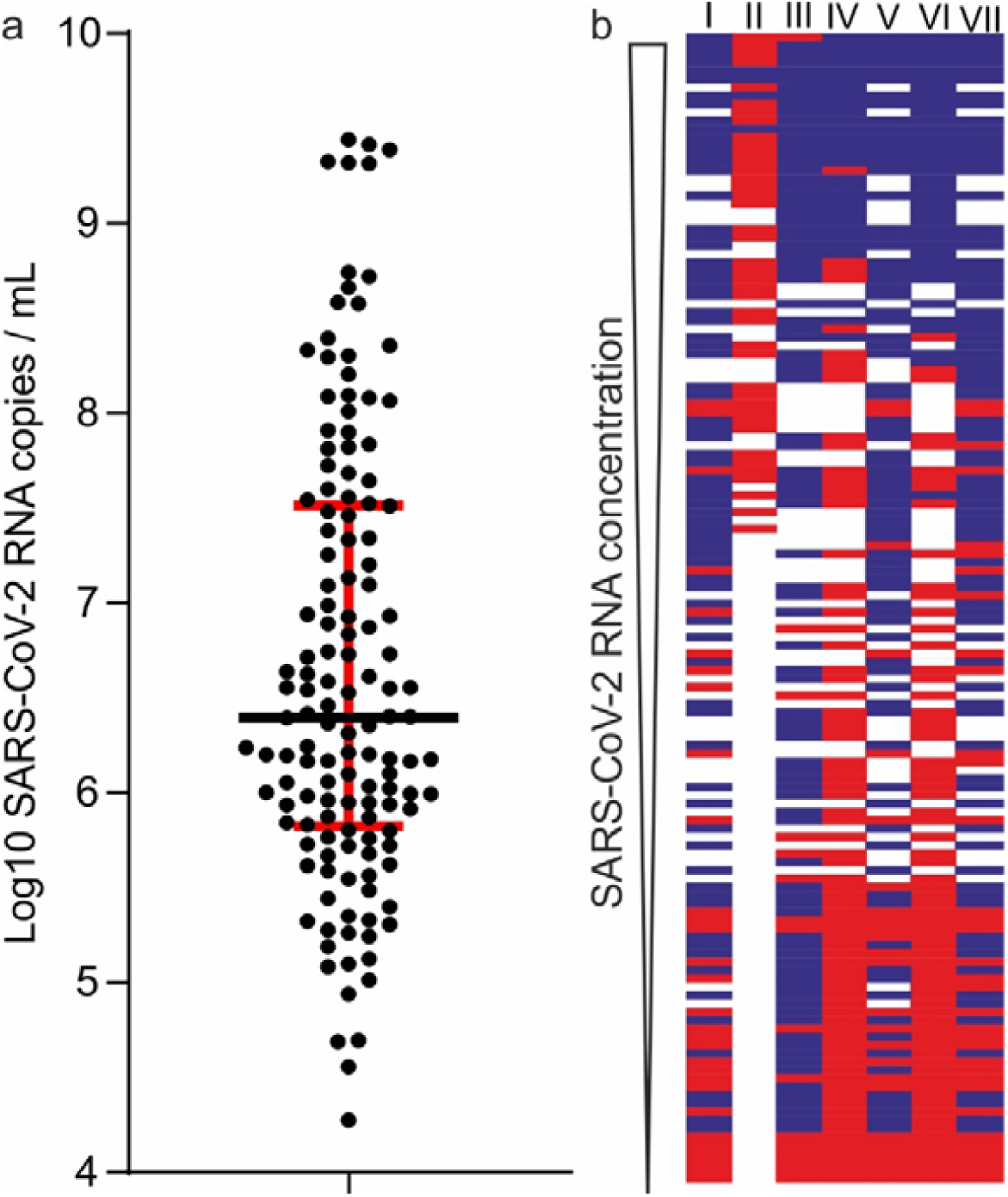
a) Distribution of SARS-CoV-2 viral RNA concentrations across clinical samples used for AgPOCT testing. b) Overview of tested samples and corresponding outcomes in the seven AgPOCT (per column). Blue fields correspond to a positive AgPOCT result, red fields to a negative result. Empty fields represent samples that were not tested in the corresponding test.

Based on this testing, a binary logistic regression analysis was performed to determine 50% and 95% limits of detection per AgPOCT (**Supplementary Figure 1**). Without correction for the lower sample input as opposed to standard AgPOCT protocols in our study, the RT-PCR-quantified virus concentrations at which 95% hit rates are achieved ranged between 3.4 × 10^6^ and 7.41 × 10^7^ copies per ml of swab suspension for the five most sensitive assays. With correction for sample input, these figures are lower by a factor of approximately 8 (**Table 2)**.

**Table 2.** Limits of detection.

| Assay<sup>a</sup> | N. of tested samples | Limit of detection<sup>b</sup><br>Log<sub>10</sub> SARS-CoV-2 RNA copies per swab |  | Adjusted limit of detection<sup>d</sup> |
| --- | --- | --- | --- | --- |
|  |  | 50% positive AgPOCT results | 95% positive AgPOCT results | 95% mean hit rate X 0.125 |
| I | 105 | 5.61 (5.27 - 5.95) | 7.45 (6.79 - 8.20) | 6.55 copies/swab |
| II<sup>c</sup> | 45 | 9.51 (8.84 - 12.26) | 11.10 (9.71 - 17.01) | 10.20 copies/swab |
| III<sup>c</sup> | 105 | 4.48 (3.41 - 5.32) | 7.27 (6.27 - 8.40) | 6.37 copies/swab |
| IV | 105 | 7.60 (7.37 - 7.82) | 8.36 (8.00 - 8.76) | 7.46 copies/swab |
| V | 105 | 5.40 (4.99 - 5.77) | 7.22 (6.57 - 7.96) | 6.32 copies/swab |
| VI | 105 | 7.19 (6.97 - 7.43) | 7.87 (7.52 - 8.23) | 6.97 copies/swab |
| VII | 115 | 5.64 (5.28 - 6.00) | 7.68 (6.96 - 8.50) | 6.78 copies/swab |

<sup>a</sup>I: Abbott Panbio™ COVID-19 Ag Rapid Test; II. RapiGEN BIOCREDIT COVID-19 Ag; III: Healgen® Coronavirus Ag Rapid Test Cassette (Swab); IV Coris Bioconcept Covid.19 Ag Respi-Strip; V: R-Biopharm RIDA®QUICK SARS-CoV-2 Antigen; VI NAL von minden; NADAL COVID19-Ag Test; VII: Roche/SD Biosensor SARS-CoV Rapid Antigen Test
<sup>b</sup>Mean concentration that yields 50% or 95% positive results according to a binary logistic regression analysis. Numbers in parenthesis denote the 95% highest posterior density interval determined by the Bayesian binary logistic regression model. Concentration per swab presumes that swabs are resuspended in 1 mL of fluid during preanalytical processes in RT-PCR used to determine viral loads.
<sup>c</sup>Model fit was suboptimal due to a large difference in the number of positive and negative test results.
<sup>d</sup>Due to a systematic preanalytical dilution factor in our AgPOCT evaluations, the projected mean concentrations at which 95% hit rate are achieved were corrected to be 8-fold (0.9 Log<sub>10</sub>) lower. This correction factor is an average over all correction factors between the actual volume input in our validation studies and the volume input as per manufacturer's instruction. Input volumes in all cases are subject to great variability due to the undefined volumes of viscous respiratory tract specimens taken up by swab sampling devices. The here-provided statistical evaluation suggests a level of precision that does not reflect the clinical reality in AgPOCT use.

### Exclusivity testing

To determine any systematic cross-reactivity with relevant viral antigens, we tested cell- or tissue culture supernatants containing known concentrations of the four endemic human coronaviruses (HCoVs) as well as MERS- and SARS-CoV, applying 50 µl of supernatant into the lysis buffer of each AgPOCT (**Table 3**). With one exception that was not reproducible, none of the assays showed cross-reactivity towards HCoVs and MERS-CoV. SARS-CoV was cross-detected by all assays. We tested 100 stored clinical samples from patients with known acute infections caused by respiratory viruses other than SARS-CoV-2, including some samples containing mycoplasma and legionella. With one exception, all assays detected either none, one, or two false positive results in 100 tests (**Table 4**). Of note, about half of all false positive results were reproducible upon re-testing of the same sample, while there was no association with any specific known pathogen in the samples. This suggests a specific factor other than the tested pathogens to cause positive signals. In 15 samples that tested false positive in total, one sample caused a positive signal in two different assays.

**Table 3.**
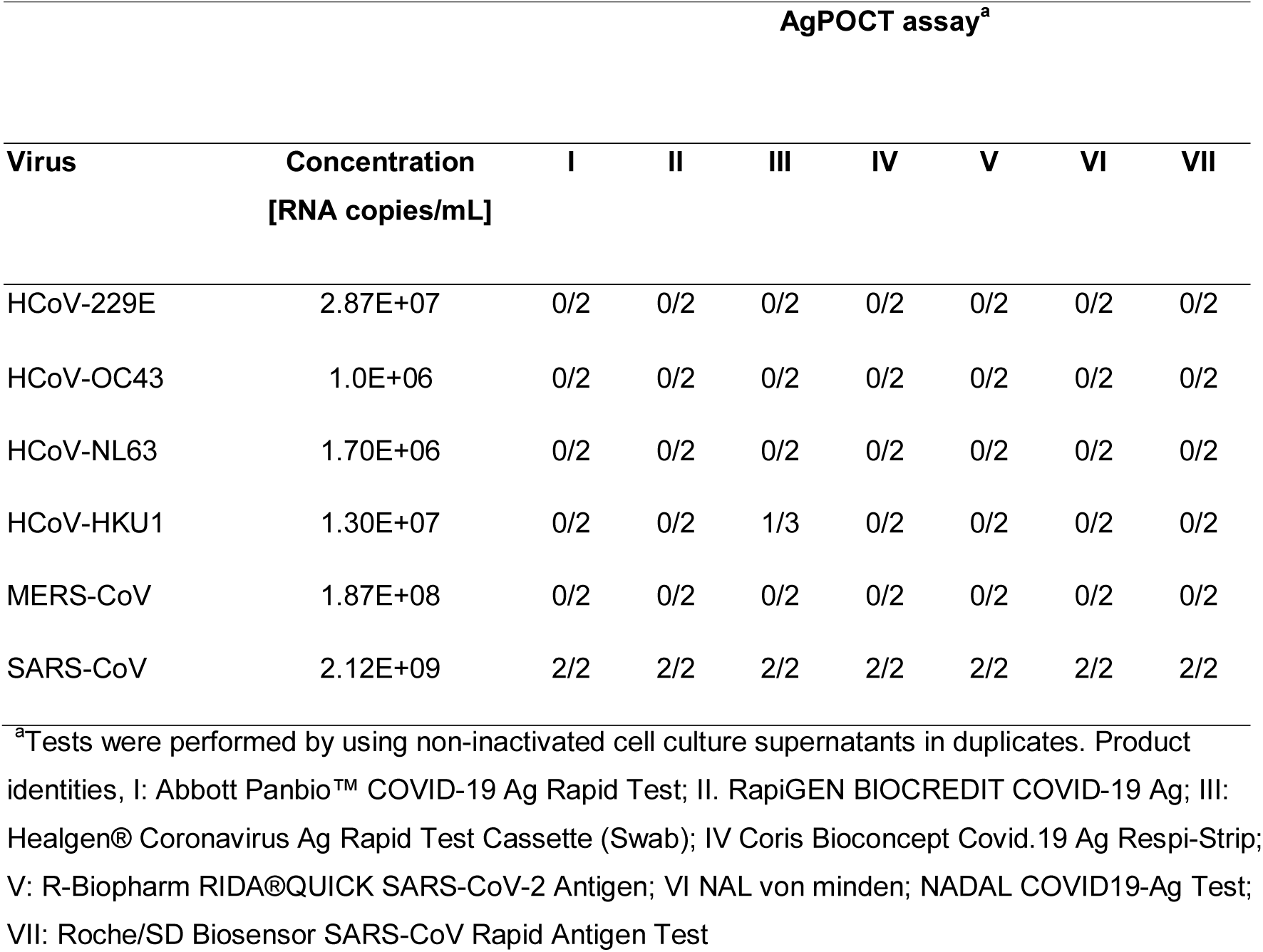
Specificity in testing using cell culture supernatants of other human coronaviruses.

**Table 4.** Specificity in the testing of clinical samples, without SARS-CoV-2 detection:

| Pathogen | N | AgPOCT assay<sup>a</sup> |  |  |  |  |  |  |
| --- | --- | --- | --- | --- | --- | --- | --- | --- |
|  |  | I | II | III | IV | V | VI | VII |
| Adenovirus | 9 | - | - | 1<sup>b</sup> | - | - | - | - |
| Bocavirus | 9 | - | - | - | - | - | - | - |
| HCoV-NL63 | 1 | - | - | - | - | - | - | - |
| HCoV-OC43 | 1 | - | - | - | - | - | - | - |
| Entero/Rhinovirus | 9 | - | - | 1<sup>b</sup> | - | - | - | - |
| Influenzavirus A H1 | 10 | - | - | 2<sup>b</sup> | - | 1<sup>c</sup> | - | - |
| Influenzavirus A H3 | 9 | - | - | 2<sup>b,c</sup> | - | 1<sup>c</sup> | - | - |
| Influenzavirus B | 1 | - | - | - | - | - | - | - |
| Metapneumovirus | 1 | - | - | - | - | - | - | - |
| Parainfluenzavirus 1 | 8 | - | - | 3<sup>b</sup> | - | - | - | - |
| Parainfluenzavirus 2 | 3 | - | - | 2<sup>b,c</sup> | - | - | - | - |
| Parainfluenzavirus 3 | 10<sup>d</sup> | - | - | 1<sup>c</sup> | - | - | - | 1<sup>b</sup> |
| RSV-A | 7 | 1<sup>b</sup> | - | - | - | - | - | - |
| RSV-B | 7 | - | - | - | - | - | - | - |
| Mycopla. pneumon. | 8 | - | - | - | - | - | - | - |
| Legion. Pneumophila | 7 | - | - | - | - | - | - | - |
| **Total** | **100** | **1** | **0** | **12** | **0** | **2** | **0** | **1** |

<sup>a</sup>I: Abbott Panbio™ COVID-19 Ag Rapid Test; II. RapiGEN BIOCREDIT COVID-19 Ag; III: Healgen® Coronavirus Ag Rapid Test Cassette (Swab); IV Coris Bioconcept Covid.19 Ag Respi-Strip; V: R-Biopharm RIDA®QUICK SARS-CoV-2 Antigen; VI NAL von minden; NADAL COVID19-Ag Test; VII: Roche/SD Biosensor SARS-CoV Rapid Antigen Test
<sup>b</sup>The non-specific positive reaction was reproduced in a repeat test.
<sup>c</sup>The non-specific positive reaction was reproduced in a repeat test in one of the two replicates.
<sup>d</sup>One of these samples also was positive in assays III and VII

### Testing of healthy volunteers

In view of the rates of false positive results in clinical samples with two of the assays, we conducted a self-testing exercise using all AgPOCT, employing healthy laboratory members without signs of respiratory tract infection. As summarized in **Table 5**, the same AgPOCT that generated false positive results with stored clinical samples also showed increased rates of positives during testing of healthy subjects. All positive results were resolved to false positive through immediate testing by RT-PCR.

**Table 5.** False positive results in 35 SARS-CoV-2 negative employees.

| AgPOCT<sup>a</sup> | I | II | III | IV | V | VI | VII |
| --- | --- | --- | --- | --- | --- | --- | --- |
| False positives | - | - | 3<sup>c</sup> | - | 5<sup>c</sup> | 1 | 1 |
| Specificity (%)<sup>b</sup> | 100 | 100 | 91.42 | 100 | 82.86 | 97.12 | 97.12 |

<sup>a</sup>I: Abbott Panbio™ COVID-19 Ag Rapid Test; II. RapiGEN BIOCREDIT COVID-19 Ag; III: Healgen® Coronavirus Ag Rapid Test Cassette (Swab); IV Coris Bioconcept Covid.19 Ag Respi-Strip; V: R-Biopharm RIDA®QUICK SARS-CoV-2 Antigen; VI NAL von minden; NADAL COVID19-Ag Test; VII: Roche/SD Biosensor SARS-CoV Rapid Antigen Test;
<sup>b</sup>In 35 subjects, 30 conducting nasopharyngeal swabs and 5 conducting pharyngeal swabs
<sup>c</sup>One person tested false positive in assays III and V

### Cumulative specificity

The cumulative specificities from exclusivity testing as well as testing of healthy volunteers were: Abbott Panbio™ COVID-19 Ag Rapid Test (99.26%), RapiGEN BIOCREDIT COVID-19 Ag (100%); Healgen® Coronavirus Ag Rapid Test Cassette (88.24%); Coris Bioconcept Covid.19 Ag Respi-Strip (100%); R-Biopharm RIDA®QUICK SARS-CoV-2 Antigen (94.85%); NAL von minden NADAL COVID19-Ag Test (99.26%); Roche/SD Biosensor SARS-CoV Rapid Antigen Test (98.53%).

## Discussion

We provide a comparison of performance of seven AgPOCT assays that have recently become available on the European market. These medical diagnostic devices are cleared in many countries for use outside the laboratory as long as testing results are supervised by medical personnel. The short turnaround time of these tests is expected to enable major changes in clinical and public health practice, given that sensitivity and specificity is sufficient. Because of the strong demand during a constantly evolving situation, the latter question has not been thoroughly clarified for most AgPOCT products.

The aim of the present study was to ease some of the challenges associated with the clinical evaluation of AgPOCTs during the present pandemic situation. As the arrival of prototype tests coincided with a time of low incidence over the summer months in the Northern hemisphere, the recruitment of freshly infected subjects for clinical evaluation has been difficult. Due to the rapid change of viral loads over the acute phase of COVID-19 illness (11, 17), AgPOCT have a narrow timeframe for their useful application that basically comprises the first week of symptoms. In view of the growing experience with RT-PCR testing during this timeframe, we aimed to mainly provide a reflection of test performance based on analytical properties, i.e., the approximate viral concentrations that can be detected by the assays as well as their propensity to generate false positive results.

In terms of sensitivity, the detection range of most tests seemed to range between one and ten million copies per swab (accounting for a systematic pre-dilution as explained above) and thus corresponds to a concentration that predicts a virus isolation success rate of ca. 20% in cell culture (11, 18, 19). In the cited studies, this level of isolation success is typically reached by the end of the first week of symptoms. He et al. have shown that this point in time also correlates with the end of factual transmissibility (17). Although many caveats remain, the point in the course of the first week of symptoms at which AgPOCT results turn negative may thus indicate the time at which infectivity resolves. In a situation marked by transition to higher incidence rates, the immediate availability of test results could enable novel public health concepts in which decisions to isolate or maintain isolation are based on infectivity testing rather than infection screening. Upon first patient contact, a positive result in AgPOCT could also help physicians decide on immediate isolation measures based on the identification of individuals who shed particularly large amounts of virus. In hospitalized patients at the end of their clinical course, negative AgPOCT results may provide an additional criterion to safely discharge patients.

Screening of asymptomatic subjects with the expectation of absence of virus is more difficult. Given the limitations of sensitivity, the results of AgPOCT should be understood as a momentary assessment of infectiousness rather than a diagnosis with power to exclude infection. As there is a steep incline of virus concentration around or before the onset of symptoms, guidelines for using AgPOCT should mention that a negative test results may reflect a lack of sensitivity, particularly when symptoms occur short after testing. Instructions that limit the validity of a negative test result in healthy subjects to the day of application could be used to address this challenge.

Also, the limited specificity of most AgPOCT should trigger RT-PCR confirmation of positive tests whenever possible. We have seen acceptable rates of false positive results with most AgPOCT but rates around 10% with two assays in particular. One of these assays (R-Biopharm) was tested here as a preliminary version predating the marketed product. The other assay may suffer from lot-to-lot variability as an independent study of the same product does not show comparable issues with false positives (information based on product insert by the distributor, HealGen).

There are clear limitations to our study. For instance, we can only provide an approximate sensitivity assessment for individual AgPOCT as we used stored samples on which we had to apply equal preanalytical treatments despite slight differences between kits in terms of the size of the swab samples. An absolute assessment of limits of detection for each test, as well as a strict comparison of relative sensitivities is therefore not possible. Also, the encountered issues with specificity of two products are likely to be transitory issues that can likely be amended by adjustments of reagent concentrations and improvements of production processes in the very near future, perhaps even before some products become widely available. Our study finally does not compare practical differences between assays, for instance, whether sample buffers come as bulk volume or are pre-filled in reaction tubes. These issues are a main subject to the qualification of products as consumer-grade tests (home tests), a process that is underway for some but not all products. There are other limitations, including the absence of clinical information due to anonymization of samples. Nevertheless, the present contribution provides an early impression about the performance of AgPOCT of several major distributors.

## Data Availability

All data are available within the manuscript.

## Acknowledgements

Parts of this work was funded by European Union DG Research through projects Prepare (GA602525) and Compare (GA643476) to CD, the German Ministry of Research through projects RAPID (01KI1723A) and DZIF (301-4-7-01.703) to CD, by the Federal Ministry for Economic Affairs and Energy (ZIM 16KN073824) to VMC, and by the German Ministry of Health (Konsiliarlabor für Coronaviren) to CD and VMC. This study is based on research funded in part by the Bill &Melinda Gates Foundation (grant ID INV-005971) to JFD and CD. The findings and conclusions contained within are those of the authors and do not necessarily reflect positions or policies of the Bill & Melinda Gates Foundation.

