## Supplementary Figure 1 for "Comparison of seven commercial SARS-CoV-2 rapid Point-of-Care Antigen tests"

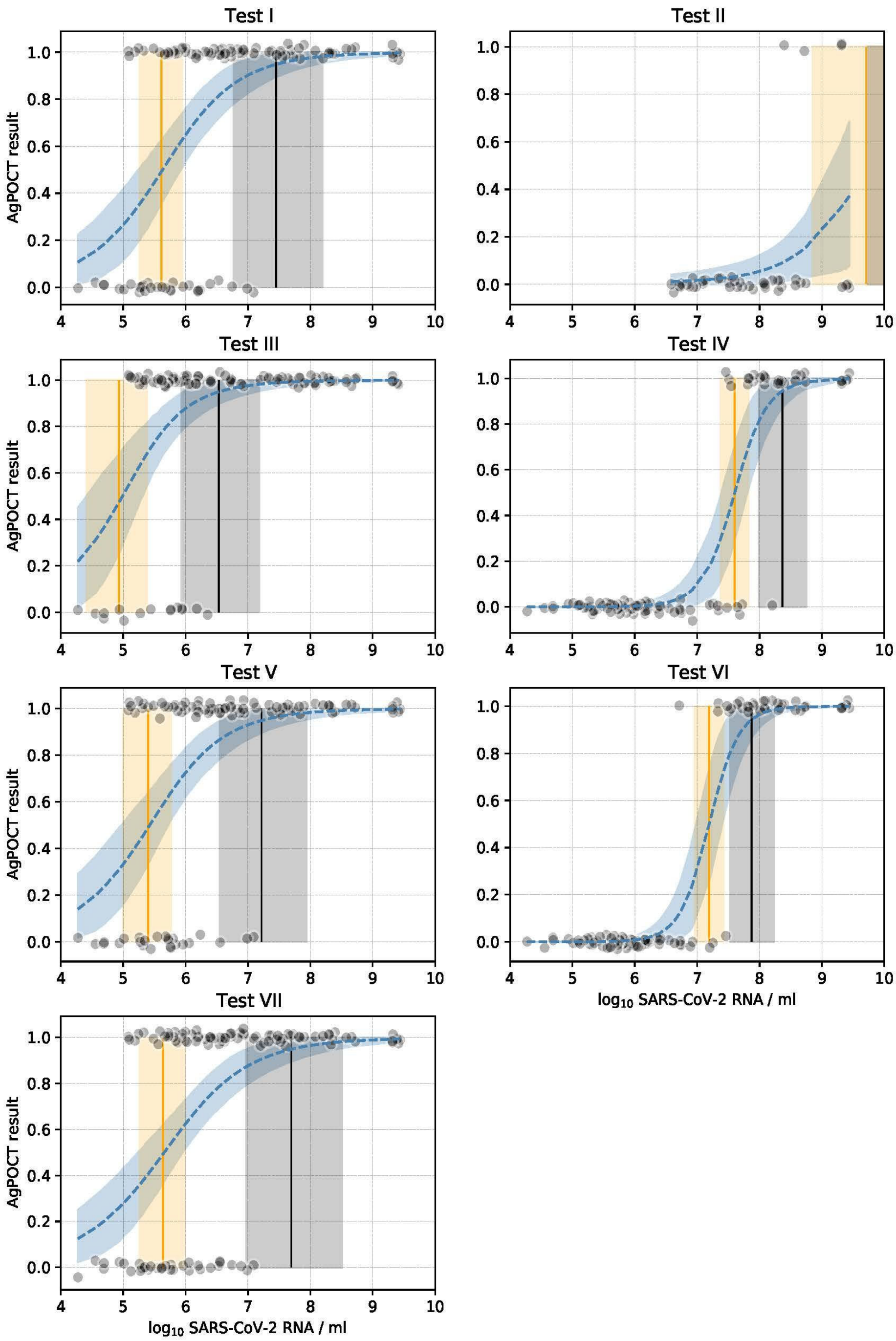


**Supplementary Figure 1:** Predicted AgPOCT results as a function of $\log_{10}$ SARS-CoV-2 RNA / mL using logistic regression. Black dots show the $\log_{10}$ SARS-CoV-2 RNA / mL (with jitter added on the y-axis) against positive (1.0) and negative (0.0) AgPOCT result. The dark blue dashed line and shaded region represent the mean and 95% highest posterior density (HPD) interval of the logistic curve fitted to the data. The orange and black vertical lines and shaded areas correspond to the mean and 95% HPD interval of the threshold log10 SARS-CoV-2 RNA / ml where 50% (orange) and 95% (black) of the AgPOCT test results have a value of 1.0, corresponding to a positive result (also see Table 1). Model fit for Test II and Test III was poor, due to a large difference in the number of positive and negative test results.
